# Deploying Smartphone-Based Artificial Intelligence Interventions to Community Health Workers in Low- and Middle-Income Countries: Recommendations for Product Developers, Implementing Partners, and Governments

**DOI:** 10.64898/2025.12.29.25343175

**Authors:** Elina Urli Hodges, Kate Crissman, Zhanyue Chang, Kiara Ekeigwe, Joao Ricardo Nickenig Vissoci, Krishna Udayakumar

## Abstract

Community health workers are crucial to public health efforts in low– and middle-income countries. Despite their contributions to improving health, they encounter numerous barriers in performing their day-to-day work. Artificial intelligence applications may offer a potential solution to some of these barriers. We examined the implementation of smartphone-based artificial intelligence interventions with community health workers in Uganda, Rwanda, and Nigeria, including how the interventions were developed, how they were used, and successes and challenges of their implementation. This research identified four key considerations for intervention developers, implementing partners, and governments, with respect to implementation of these applications, including: 1) empowering community health workers through professionalization, compensation, and training; 2) understanding the digital ecosystem and aligning with digitization efforts; 3) designing the solution to fit local context, ensuring the development and training of the intervention is applicable to the local environment; and, 4) managing data responsibly with adherence to data privacy and security regulations. This research describes the opportunities for filling gaps in supervisory support, improving diagnosis, and making work more efficient. Providing the community health workforce with digital tools replaces onerous, inefficient paper-based record keeping, enables opportunities for improved supervision, and facilitates decision-making in settings where these workers may be the only point of access to health services and information.

**Author Summary:** Community health workers are often the backbone of the health workforce in low– and middle-income countries. Despite their contributions to community health improvement, they often face barriers and inefficiencies in delivering services. The expansion of smartphone-based artificial intelligence interventions may offer solutions. We examined the implementation of smartphone-based artificial intelligence interventions with community health workers in Uganda, Rwanda, and Nigeria. We found that smartphone-based artificial intelligence applications may help to replace onerous, inefficient paper-based record keeping, enable opportunities for improved supervision, and facilitate decision-making in settings where these individuals may be the only health service providers. This research also identified four key considerations for intervention developers, implementing partners, and governments, with respect to implementation of these applications, including: 1) empowering community health workers through professionalization, compensation, and training; 2) understanding the digital ecosystem and aligning with digitization efforts; 3) designing the solution to fit local context, ensuring the development and training of the intervention is applicable to the local environment; and, 4) managing data responsibly with adherence to data privacy and security regulations.

## Introduction

As the core workforce of many public health efforts in low– and middle-income countries (LMICs), community health workers (CHWs), typically individuals without formal medical training, are at the forefront of providing access to health services, particularly among vulnerable and last-mile, rural populations [1]. This cadre of health worker is important, helping fill the gap from shortages of nurses and doctors in LMICs while advancing new, decentralized models of care, and in their role in supporting efforts towards the third Sustainable Development Goal focused on achieving good health and well-being worldwide [1, 2, 3, 4]. CHWs are often relied upon in LMICs to fill health care gaps, but face many challenges including their remuneration, lack of funding for programs, and inconsistent integration into health system structures and policies [5].

The growth of digital health technologies may offer a mechanism to alleviate some of these challenges in health delivery by CHWs [6, 7, 8]. Recent rapid growth and uptake of artificial intelligence (AI), as a type of digital technology, has increased global attention on its role in supporting health service delivery, highlighting a need for additional research on AI and health, and more specifically AI and CHWs in their role in health service delivery in low-resource, last-mile populations [9].

AI is not a new concept, and has been growing and evolving since it was formally defined in 1956 [10]. AI can be broadly defined as “any technique that enables computers to mimic human-like intelligence” [11]. AI requires data to learn from, in order to mimic this intelligence and solve problems. Algorithms, or sets of rules and instructions, guide a computer’s learning and tell it what problem to solve. The computer can run the algorithms based on data and be completely overseen by humans (supervised) or can execute algorithms without human guidance (unsupervised), or in a semi-structured way with some human oversight. Supervised algorithms use already labeled data (raw data that has been annotated with contextually relevant labels) that the AI uses as guidelines for its learning, whereas unsupervised algorithms enable the AI to learn on its own without the boundaries set by already labeled data [12].

A large portion of AI research and applications are focused in high-income countries, but there has been increased and ongoing dialogue on the potential for AI in LMICs to address health challenges [11, 13]. In its 2020 report, *Reimagining Global Health through Artificial Intelligence: The Roadmap to AI Maturity*, the Broadband Commission for Sustainable Development Working Group on Digital and AI in Health summarizes key global health issues that have the potential to be addressed by AI including health worker shortage; emerging health threats; dual burden of disease; underserved populations; the widening of health inequities as a result of urbanization; and misinformation and disinformation [11]. While the potential impact of AI in global health is highlighted, it is also noted that there is a need for more common frameworks and understanding of AI, supported by more research, for sustainable, ethical, and responsible implementation [12, 14, 15, 16].

In order to add to the evidence base around AI use in LMICs, we systematically identified three examples of how AI has been used by CHWs in three African countries. We explored how the AI interventions were developed, how they were used by CHWs, and successes and challenges of their implementation.

## Results

### Use Case Selection

From the original search processes, we identified 170 AI interventions. Upon further review and application of the selection criteria, we narrowed the list to 22 interventions (see Appendix 2). In the process of following up with the 22 intervention developers, two had no contact information, three did not clearly fit the selection criteria, nine did not respond to our outreach, and of the eight that responded, we selected three that fit all three criteria, had the most promising evidence of impact, and had developers interested in participating further in the research process. Table 1 provides an overview of the three use cases and Appendix 3 offers a more detailed description of each use case.

**Table 1.** Use case overviews.

| Table 1. Use case overviews |  |  |  |
| --- | --- | --- | --- |
|  | **Use Case 1: Appropriately identifying individuals to support CHW health service delivery** | **Use Case 2: Improving malaria diagnosis by CHWs** | **Use Case 3: Strengthening CHW adherence to clinical guidelines** |
| What is the problem the AI solution is addressing in this use case? | Tackling the problem of effective identification of individuals—time spent trying to identify an individual and link them with their medical record reduces time available for the CHW to see more patients. | CHWs handle over 60% of uncomplicated malaria cases in Rwanda, but management and support for CHWs is insufficient. | CHWs typically receive limited training that focuses on specific areas like maternal and child health. They do not receive general healthcare capacity building training for the broader population, and therefore are unable to provide integrated comprehensive health services that adhere at a high level to clinical guidelines, such as the integrated management of childhood illness (IMCI), for the assessment, diagnosis and treatment of patients. |
| Geography of implementation | Buikwe District, Uganda | Gicumbi, Nyaruguru, Nyamagabe districts, Rwanda | Kano State, Nigeria |
| AI process implemented (according to GSMA categories) | BIOMETRICS: AI-enabled facial biometrics (to identify faces for patient record identification and linking) | COMPUTER VISION: Supervised machine learning (interpreting rapid diagnostic test (RDT) results) and computer vision algorithms (identifying/analyzing uploaded images of RDTs) | DECISION MANAGEMENT: Supervised machine learning with Bayesian weighted algorithms to provide a clinical risk assessment |
| AI use case (according to Broadband Commission report categories) | OPTIMIZATION OF HEALTH OPERATIONS: integration into clinical workflow as a means to streamline patient identification | POPULATION HEALTH: solution to monitor/assess the health of human population<br>OPTIMIZATION OF HEALTH OPERATIONS: disease monitoring and surveillance, data accuracy | CLINICAL CARE PATHWAYS: Clinical decision support through analysis of meta and clinical data inputs<br>OPTIMIZATION OF HEALTH OPERATIONS: Improving healthcare capacity with quality clinical disease risk assessments and care plan generation to support app improvement, localization of recommendations, and sensitivity and specificity of clinical risk assessment |
| Scope of implementation | Research study<br>Objective: To improve and strengthen the capacity of Community Health Workers in rural Uganda for effective delivery of the COVID19 vaccine, | Research study<br>Objective: Determine feasibility and effectiveness of introducing a customized mobile app for CHWs as a quality control and scaled supervision tool to identify where | Research study<br>Objective: To develop new machine learning based clinical risk assessment logic using diagnostic truth data, and measure utilization of the |
|  | and to support the foundation for future scaling across the existing routine immunisation portfolio<br><u>Funder:</u> Corporate foundation<br><u>Implementation:</u> 200 CHWs (goal)<br><u>Study timeframe:</u> July 2023 – current | additional training and technical assistance was needed<br><u>Funder:</u> Internal funding and family foundation<br><u>Implementation:</u> 200 CHWs from six facilities in 3 high malaria burden districts in Rwanda<br><u>Study timeframe:</u> Spring 2023 | THINKMD clinical decision support tool, including usability and accessibility, to understand if it increases adherence to WHO-IMCI guidelines, particularly for danger signs in children under five by CHWs<br><u>Funding:</u> THINKMD & Kano State Primary Health Care Management Board<br><u>Implementation:</u> 15 health facilities in Kano State, Nigeria<br><u>Study timeframe:</u> April to August 2018 |
| Evidence of Potential Impact | CHW survey results (n=10) from pre- and post-biometrics application implementation:<br>The AI application helps to eliminate duplicates: 30% response pre-training to 70% response post-training<br>Saves time to identify patients: 0% response pre-training to 80% response post training<br>*(Mobiklinic & Simprints Technology. Integration of biometrics in a digital community health system - user survey finding. August 2023.)* | CHW knowledge of time required to read test result after adding sample and buffer: 79.9% (pre-intervention) and 97% (post-intervention)<br>CHW identification of positive results from faint visible lines: 13% (pre) and 85.2% (post)<br>93% of CHWs indicated the app was easy to use, 99% that the app instructions were clear and easy to follow, and 93.8% said that using the app saved time compared to paper registers<br>AI performance exceeded CHW performance compared to reference panel (target: 90%): 95.5% (AI) compared to 94% (CHWs)<br>AI identified 9% of CHWs who may need additional training or support<br>*(Umulisa N., Mwiseneza E., Maricich N., et al. Digitally-Enhanced Rapid Malaria Testing Using Artificial Intelligence (AI) to Support Quality Control with Community Health Workers in Rwanda. Poster presentation at the American Society of Tropical Medicine & Hygiene. 2023.)* | 50% actual improvement in ability of CHW to appropriately diagnose sick children, according to the WHO-IMCI-guidelines using THINKMD technology versus the typical paper-based approach<br>93% of CHWs said technology was easy to learn and use 100% said the technology was a valuable training tool<br>100% said the technology helped them identify sick children, and 100% said they were extremely likely or likely to recommend use of the technology<br>*(THINKMD. Data-driven decision support to improve community-based diagnosis and treatment. Use case. PATH/Digital Square. 2020. https://thinkmd.org/path-digital-square-2020-data-driven-decision-support-to-improve-community-based-diagnosis-and-treatment/ last accessed November 2023.* |
| Developer & Implementer | Simprints (nonprofit, developer)<br>Mobiklinic (nonprofit, implementer) | Audere (nonprofit, developer)<br>Jhpiego (nonprofit, implementer) | THINKMD (for profit, developer)<br>eHealth Africa (nonprofit, implementer) |

The first example from Uganda, highlights how facial biometrics, a type of AI, can support CHWs as they seek to enroll new patients into health services or identify existing patients and link them with their health records. The second example from Rwanda, describes the use of a type of AI called computer vision to read the results of rapid diagnostic tests for malaria, to address the challenge of CHWs who may misinterpret the test results. The third example from Nigeria, uses a type of machine learning process called clinical logic development to support CHWs in assessing at-risk children for various diseases.

All three use cases were implemented as research studies or pilots, indicating the limited scale-up of these approaches. Additionally, two of the three examples were implemented in 2023, with one ongoing, highlighting potentially a nascence of the use of these types of interventions in these settings. From the interviews with the developers and implementing partners of these use cases, we identified the following enablers for more effective implementation of an AI-powered solution with CHWs:

**Table 2.**
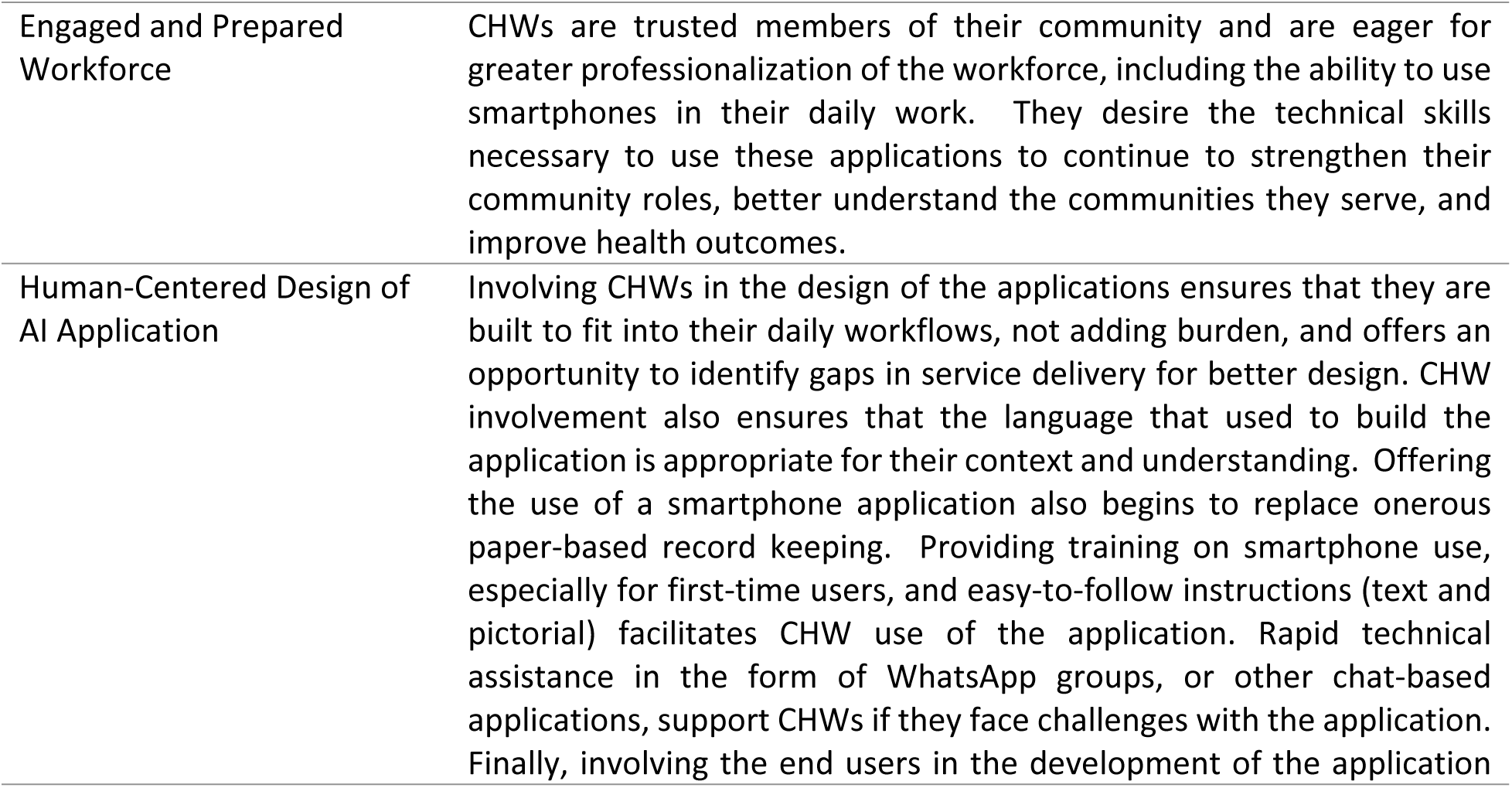
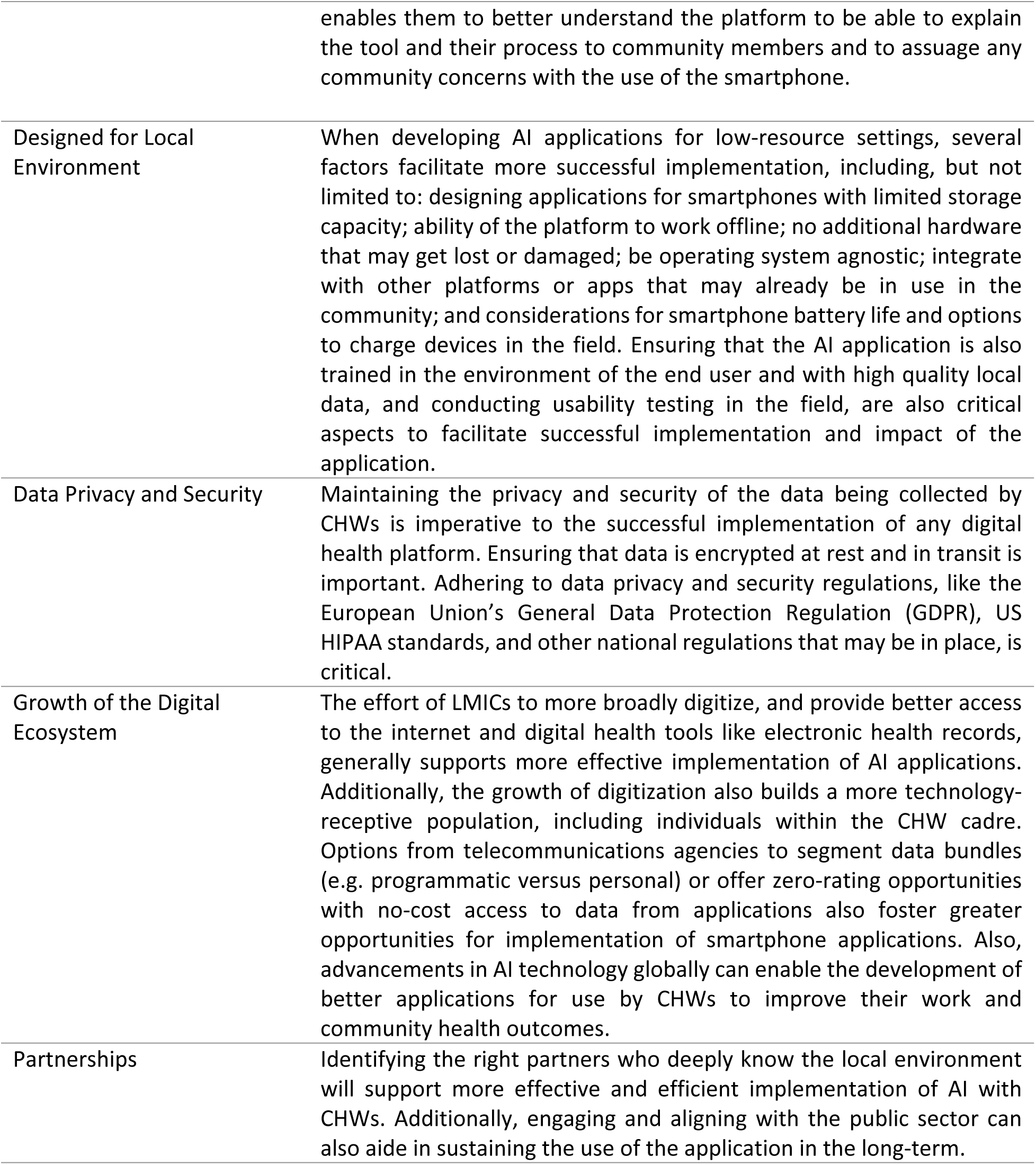
Enablers of More Effective Implementation of AI with CHWs.

|  |  |
| --- | --- |
| Engaged and Prepared Workforce | CHWs are trusted members of their community and are eager for greater professionalization of the workforce, including the ability to use smartphones in their daily work. They desire the technical skills necessary to use these applications to continue to strengthen their community roles, better understand the communities they serve, and improve health outcomes. |
| Human-Centered Design of AI Application | Involving CHWs in the design of the applications ensures that they are built to fit into their daily workflows, not adding burden, and offers an opportunity to identify gaps in service delivery for better design. CHW involvement also ensures that the language that used to build the application is appropriate for their context and understanding. Offering the use of a smartphone application also begins to replace onerous paper-based record keeping. Providing training on smartphone use, especially for first-time users, and easy-to-follow instructions (text and pictorial) facilitates CHW use of the application. Rapid technical assistance in the form of WhatsApp groups, or other chat-based applications, support CHWs if they face challenges with the application. Finally, involving the end users in the development of the application |
|  | enables them to better understand the platform to be able to explain the tool and their process to community members and to assuage any community concerns with the use of the smartphone. |
| Designed for Local Environment | When developing AI applications for low-resource settings, several factors facilitate more successful implementation, including, but not limited to: designing applications for smartphones with limited storage capacity; ability of the platform to work offline; no additional hardware that may get lost or damaged; be operating system agnostic; integrate with other platforms or apps that may already be in use in the community; and considerations for smartphone battery life and options to charge devices in the field. Ensuring that the AI application is also trained in the environment of the end user and with high quality local data, and conducting usability testing in the field, are also critical aspects to facilitate successful implementation and impact of the application. |
| Data Privacy and Security | Maintaining the privacy and security of the data being collected by CHWs is imperative to the successful implementation of any digital health platform. Ensuring that data is encrypted at rest and in transit is important. Adhering to data privacy and security regulations, like the European Union's General Data Protection Regulation (GDPR), US HIPAA standards, and other national regulations that may be in place, is critical. |
| Growth of the Digital Ecosystem | The effort of LMICs to more broadly digitize, and provide better access to the internet and digital health tools like electronic health records, generally supports more effective implementation of AI applications. Additionally, the growth of digitization also builds a more technology-receptive population, including individuals within the CHW cadre. Options from telecommunications agencies to segment data bundles (e.g. programmatic versus personal) or offer zero-rating opportunities with no-cost access to data from applications also foster greater opportunities for implementation of smartphone applications. Also, advancements in AI technology globally can enable the development of better applications for use by CHWs to improve their work and community health outcomes. |
| Partnerships | Identifying the right partners who deeply know the local environment will support more effective and efficient implementation of AI with CHWs. Additionally, engaging and aligning with the public sector can also aide in sustaining the use of the application in the long-term. |

### Recommendations

In assessing these enablers of implementation across the three use cases, we identified the following recommendations, in priority order, for AI developers, implementing partners at the local level, and governments intending to implement an AI solution with CHWs in an LMIC. The thematic areas are in alignment with the six areas for sustainable and mature AI implementation, as espoused by the Broadband Commission: People and workforce; Data and technology; Governance and regulatory; Design and process; Partnerships and stakeholders; and Business models.

1. **ENABLE THE WORKFORCE.** CHWs in LMICs are a vital cadre of the health workforce, and while their scope of work may be more limited, they are a trusted member of the healthcare team and **governments** should professionalize and remunerate this workforce. CHWs may not be as technologically savvy or may not have had prior access to smartphones on which these interventions are based, so **developers and implementers** should provide appropriate training (considering reading and technology literacy levels of the participants) on how to use a smart phone, including turning it on, navigating to apps, using apps, taking and uploading photos, and understanding how to use data. Importantly, **implementers** should institute feedback loops in which the CHWs are collecting community health data and, subsequently, any analyses from the collected data are provided back to the CHWs, so that they can see how their data is being used and have greater awareness of community trends.
2. **UNDERSTAND THE ECOSYSTEM.** Taking time to understand the digital ecosystem in the setting into which your intervention may be used may facilitate faster uptake. **Governments** should support wider digitization efforts to see greater impact from and access to AI interventions. Additionally, **developers** should work with the right partners with a deep understanding of the local health context and workforce to implement the intervention, to drive greater reach, impact, and potentially sustain the efforts, particularly those that are connected with the public sector. **Developers and implementers** should work collaboratively to identify the right telecommunications company to provide internet services depending on the location of implementation. Finally, **developers** offering AI solutions to CHW communities should understand that subscription-based contracts may be less appealing to entities involved in community health work as the workload with CHWs is not necessarily consistent or predictable; developers may wish to consider per patient or per transaction fees for use of their application, in order to be more competitive.
3. **DESIGN THE AI SOLUTION TO FIT LOCAL CONTEXT.** The recommendations in this category are primarily for the **developers** of AI solutions interested in making their applications available in LMICs. The recommendations are bucketed into three categories: a) development and training of the AI intervention; b) smartphone integration and costs; and c) end user applicability.
  a) <u>Development and training of AI intervention.</u> Co-design the AI solution based on solving local challenges. Take the time to understand the problems on the ground by working with the Ministry of Health and/or local health system leaders to determine if AI is even necessary, and if so, this engage will support the development of the most appropriate solution. Consider that there may be bias in the data that you are training your AI on, in particular if applying the solution to LMIC settings; large data sets are primarily from high income settings like the United States or Europe. Implement robust real-world, high quality data collection from the local settings into which you are planning to implement the intervention to limit bias and ensure a focus on the end user’s environment. Additionally, if building an AI intervention for frontline health workers, consider that medical terminology may be different depending on context and therefore implement an iterative learning process to ensure that the language used in the intervention is the same as that in the field. Finally, build the intervention to collect only the data that is needed to train the model, additional data may not be needed and may even cause added security risks for individuals, such as personal location information.
  b) <u>Smartphone integration and costs.</u> As with many digital health interventions using a smartphone in the field, consider that older phones may not have much storage capacity for larger files such as images, internet is likely not available so building for an offline environment may be critical, and battery life is essential. **Implementers** in particular may want to consider equipping CHWs with portable/solar phone chargers or find other power sources. As one CHW noted, “when you wake up without battery, that means you are not going to work that day.” There may be costs associated with storing images on phones and uploading images to cloud servers, or on to servers based in countries, so considerations must be made for how data use costs will be handled. If applicable, consider purchasing data bundles that can segment out which costs are programmatic and which are personal or explore zero-rating opportunities (no-cost data traffic when using the application) with a local telecommunications company. Budget considerations must also be made for the purchase of smartphones on which the applications are integrated. It is also important to consider building an intervention that is operating system agnostic (e.g. iOS, Android) and that there is a consideration for the language used in the operating system itself, as English is often not prevalent in these last mile settings.
  c) <u>End user applicability.</u> Developers must ensure that the AI intervention fits seamlessly into the workflow of the CHWs to ensure the greatest efficiency and impact, and to not be a burden to either CHW or beneficiary (patient) during use. Conducting usability testing together with partners in the field will be critical to ensure fit to workflow. Supporting this workforce with easy-to-follow pictorial instructions, displaying content in their local language, and providing troubleshooting support through platforms such as WhatsApp, will facilitate use of the intervention.
4. **BE RESPONSIBLE WITH DATA.** Data is a prerequisite for any AI intervention. **All stakeholders** involved in the deployment of AI interventions with CHWs must be aware of data privacy and security regulations. Examples of data regulations include the GDPR which imposes requirements on any organization in the world that is collecting data relating to European Union citizens, Nigeria’s recently updated Data Protection Act of 2023 which includes aspects of artificial intelligence, South Africa’s Protection of Personal Information Act (PoPIA) policy, and the East African regional Intergovernmental Authority on Development’s recently adopted Regional Health Data Sharing and Protection Policy Framework [17]. Regular education of all stakeholders on updated regulations is imperative, with recognition that compliance with regularly changing regulations and a lack of a broader ethical framework for country-to-country localizations may be challenging for developers.

Health data is particularly sensitive, depending on the type of data being collected, and as stated previously, considerations must be made to collect only the data that is required for continuous validation and improvement. **Developers** should ensure that data is encrypted at rest and in transit to storage. **Developers**, **in partnership with national and global entities,** may also wish to generate greater awareness of AI and what it will take to ensure data security and privacy, and what it will take to maintain and improve the accuracy of these interventions for the benefit of the target population of users or beneficiaries. **Implementers** working with CHWs must offer training on the sensitivity of the data that exists in the phones and AI applications themselves, to ensure that CHWs understand the importance of maintaining the confidentiality of the information and securing the devices. **Developers and governments**, in settings where policy and regulatory systems are in place and implemented to protect data privacy and security, may consider negotiating access to data by governments.

## Discussion

Given the significant rise of the use of AI in health settings and the limited data coming from LMICs on the topic, to the best of our knowledge, this paper is the first attempt to identify considerations for implementing artificial intelligence with the CHW workforce in LMICs. While much of the world is focused on the rise of large language models like ChatGPT, and other AI models whose learning is not supervised by humans, the AI models presented in these use cases are regularly reviewed and assessed for their accuracy and calibrated to ensure that the machines are not independently learning from the inputted data without a human in the loop.

Faced with a myriad of challenges in delivering health services to last mile populations, CHWs equipped and empowered with digital tools like smartphone-based applications may be more efficient and effective in delivering services. Using AI to predict which CHWs need the most support in performing services may enable more targeted supervision and training opportunities, with limited human resources. Process optimization tools like those described in the use cases presented may save CHW time and increase operational efficiencies; they have been recommended as a starting point for health system leaders seeking to implement AI interventions [18]. A single solution may also incorporate multiple forms of AI, thereby simplifying and minimizing the number of applications that a CHW may need to use.

While AI is likely to become more ubiquitous in health, LMIC settings may pose unique implementation challenges given the array of considerations that must be made to ensure successful scaling, including sufficient regulatory and data governance policies, regular internet access, data availability and financing [18]. Additionally, there may be challenges unique to the AI developers themselves and their business models, in particular those who need to train models on high quality data from the specific local environment – necessitating potentially a model for each unique country or setting or population. Conversely, LMIC governments and health system leaders have the opportunity to leapfrog and embrace digital health trends, including AI, that can overcome human resource, education, and service quality challenges in particular.

We recognize the limitations from this exercise, particularly that this is not a comprehensive study of the AI interventions being deployed with CHWs, as there may be others we were not able to identify through our search process. Additionally, the impact from these interventions is self-reported and do not include clinical or operational outcomes; more independent research must be done to gauge both process and outcomes impact. There are also very few long-established use cases of the implementation of AI in LMIC settings.

Recent research has shown that AI deployed through smartphones can be used by CHWs with proper training and resources, and integration into the health system [19]. This research describes the opportunities for filling gaps in supervisory support, improving diagnosis, and making the work of CHWs more efficient. Providing CHWs with digital tools replaces onerous, inefficient paper-based record keeping, enables opportunities for improved supervision, and facilitates decision-making in settings where these workers may be the only point of access to health services and information. Additional research in this field is needed to measure the impact and cost of these technologies, including the cost of data storage and internet data usage fees, and to better understand how to scale and sustain these tools in light of their potential for health systems workforce strengthening.

## Methods

### Use Case Selection

We developed three criteria to select the AI interventions from which we would develop the use cases: 1) the intervention must be operating in an LMIC, 2) the intervention deployed must be AI in some form, and, 3) the intervention must be used by, or in support of, CHWs. Additionally, we looked for any description of potential impact of the interventions. We took three search approaches to identify the interventions: 1) we conducted a Google search with the search terms: AI OR Artificial Intelligence and Community Health Workers OR CHWs OR CHW OR Community Health Volunteers OR CHVs OR CHV and LMIC OR Low– and middle-income country OR developing country OR LDC OR least developed country and social enterprise OR nonprofit OR non-governmental organization OR NGO OR innovator OR innovation; 2) we surveyed the Innovations in Healthcare Innovator Network for referrals; and, 3) we reviewed every AI intervention for health listed in the aforementioned Broadband Commission report, the GSMA report, and the USAID report [11, 12, 14].

Once these activities were completed, we reviewed each intervention identified through this process, including a review of websites, documents, and other information provided by the developer, and applied the selection criteria to narrow the list. Once the list was narrowed, we outreached to the developer of each AI intervention to request additional information on the type of AI deployed, how the intervention was being deployed to CHWs (confirming its use with our definition of a CHW, not another clinical cadre), and other questions to assist us in confirming the fit of the intervention to the selection criteria. Upon another review of the information provided to us by the developers, we narrowed the list further and selected the final three interventions, and their corresponding use cases, based upon the selection criteria, available evidence of impact, and their level of engagement with us.

### Semi-Structured Key Informant Interviews

We conducted semi-structured key informant interviews with the developers of the three AI interventions and their partners located in the countries where the interventions were implemented for the use cases. We developed an interview guide (Appendix 1) to inform our understanding of how the AI interventions were used with CHWs and the enablers and barriers to their implementation. The interview guide was informed by the Broadband Commission report’s six areas of AI maturity (People and workforce; Data and technology; Governance and regulatory; Design and process; Partnerships and stakeholders; and Business models) which are key characteristics for building a more sustainable and mature environment for AI implementation. We also included questions on the enablers and barriers of implementation, and questions focused on impact and recommendations for the field. Interview findings were summarized against these thematic areas and incorporated into case studies (Appendix 3), with a summary of enablers or considerations of deployment of AI interventions to CHWs, across all three use cases, presented in the following Results section.

### Document and Literature Review

In addition to interviews, we reviewed the intervention developers’ websites, including reports, conference abstracts, case studies, and other materials, and reviewed any journal articles that published their activities and impact, to identify additional insights regarding technology development as well as implementation.

This research (protocol 2024-0054) received a waiver for exemption from the Duke University Institutional Review Board as it does not qualify as research with human subjects.

## Data Availability

All relevant data are within the manuscript and its Supporting Information files.

## Acknowledgments

The authors would like to thank the teams from Audere, Simprints, THINKMD, MobiKlinic, Jhpiego Rwanda, and E-Health Africa who reviewed this paper, and offered their insights and access to data to inform this paper.

## Study Funding

This study was funded by the Bayer Cares Foundation.

## APC Funding

The article processing charge was funded by Innovations in Healthcare

## Notes

### Competing Interest Statement

The authors have declared no competing interest.

### Funding Statement

Yes

